# Acute kidney injury after cardiac surgery is associated with platelet activation

**DOI:** 10.1101/2023.10.10.23296815

**Authors:** Naomi Brown, Nikol Sullo, Nathan Tyson, Bryony Eagle-Hemming, Florence Y. Lai, Sophia Sheikh, Kristina Tomkova, Lathishia Joel-David, Tracy Kumar, Hardeep Aujla, Alison H Goodall, Gavin J Murphy, Marcin J Woźniak

## Abstract

**Background:** Post-cardiac surgery acute kidney injury (AKI) is common, with high rates of mortality and morbidity. Despite extensive research, the AKI pathogenesis is still unknown. We hypothesised that platelet activation, circulating extracellular vesicles (EV) and micro-RNA levels associate with post-surgery AKI.

**Methods:** Plasma samples were collected from 95 patients recruited to the MaRACAS study before, immediately after and 6-12, 24 and 48 hours after cardiac surgery. Platelet responsiveness was measured with Multiplate. Flow cytometry was used to measure platelet and leukocyte activation and EV derivation, while EV size distribution and concentrations were measured using NanoSight. Circulating soluble biomarkers were measured by immunoassays. Micro-RNA was measured by TaqMan arrays, validated by qPCR.

**Results:** In the 57% (54/95) of patients who developed AKI post-surgery, the numbers of platelet-derived EVs were higher 24 hours after surgery. Platelets in AKI patients were desensitised to ADP 6-12 hours after surgery, independent of the administration of aspirin or P2Y12 antagonists. AKI patients also had more activated platelets 6-12 hours after surgery, more circulating platelet-granulocyte aggregates before and 6-12 and 24 hours after surgery and higher levels of sICAM1 before and 48 hours after surgery. TaqMan arrays identified miR-668 downregulated before and miR-92a-1, –920, –518a-3p, –133b and –1262 upregulated after surgery in AKI patients. qRT-PCR confirmed these differences for miR-1262.

**Conclusions:** AKI is associated with increased platelet activation during cardiac surgery, indicating that alternative platelet inhibition treatments may be renoprotective. Studies in larger cohorts are required to validate these findings.

## Introduction

Acute kidney injury (AKI) is a common complication of cardiac surgery that is associated with increased morbidity and mortality, [1] prolonged hospital stays and increases healthcare costs. [2] AKI is defined clinically by an acute decrease in glomerular filtration rate (GFR) as determined by changes in serum creatinine (SCr) concentrations and urine output. The mechanisms that underlie AKI are poorly understood, but inflammation and activation of the vascular endothelium and platelets are thought to contribute. [3]

Leukocytes and platelets play important roles in AKI pathophysiology, and their activation could serve as an AKI biomarker. Leukocytes contribute to the inflammatory response observed in AKI by releasing pro-inflammatory cytokines and reactive oxygen species. Platelet activation and aggregation within the renal microvasculature can lead to capillary occlusion, impairing blood flow and worsening tissue injury. [4] In inflammatory states, activated platelets bind monocytes and neutrophils through interaction of P-selectin with P-selectin glycoprotein ligand 1 (PSGL-1). Chemokines and cytokines released by platelets further modulate leukocyte activity, triggering neutrophil extracellular traps formation and enhancing the inflammatory response. [5,6] Extracellular vesicles (EVs) are small, membrane-bound particles secreted by cells into the extracellular space and involved in the transfer of proteins and genetic information (mRNA and microRNA) specific to their cellular origin. Micro-RNA (miRNA) are short, non-coding RNA oligonucleotides actively or passively (e.g. dead cells) secreted either within EVs, complexed with proteins (AGO-2 or NPM1 [7]) or high-density lipoproteins. [8,9] Several miRNAs have been associated with the pathogenesis of AKI, [10,11] and plasma miRNAs have been proposed as novel biomarkers for AKI in both adult [12] and paediatric [13] populations. Hence, changes in the concentration of EVs and miRNA during pathological processes can both promote pathological changes and serve as an indicator for affected tissues or cells. [14]

In acute kidney injury, platelet-derived EVs that carry pro-coagulant factors, such as tissue factor and P-selectin, can contribute to microvascular thrombosis within the kidney. [15] Platelet-derived EVs can transfer inflammatory mediators and microRNAs, like miR-223, that enhance the inflammatory response and leukocyte recruitment to the site of injury, leading to further tissue damage. [16] The interaction between platelet EVs and endothelial cells can also disrupt endothelial integrity, increasing vascular permeability and promoting injury. [17,18] Therefore, platelets and platelet-derived EVs could play a causal role in AKI and be potential therapeutic targets.

We hypothesised that platelet activation, including circulating extracellular vesicles (EV) derived from different cell types and micro-RNA levels, associate with post-surgery AKI. To test this hypothesis, we performed longitudinal measurements from baseline to 48 hours of platelet, leukocyte, and endothelial activation simultaneously with assessments of EV and miRNA in plasma from adults undergoing cardiac surgery.

## Methods

### Study design and setting

An Observational Case Control Study to Identify the Role of MV and MV Derived Micro-RNA in Post CArdiac Surgery AKI (MaRACAS) was designed as a prospective, single-centre observational study approved by East Midlands – Leicester South Research Ethics Committee (REC reference 13/EM/0383) conducted between 11^th^ December 2014 and 30^th^ May 2017 in Glenfield General Hospital in Leicester, UK. The protocol was registered at https://clinicaltrials.gov/ct2/show/NCT02315183.

### Study cohort

included prospectively recruited 95 consecutive adult cardiac surgery patients (>16 years old) undergoing coronary artery bypass grafting or valve surgery with moderately hypothermic cardiopulmonary bypass (32 – 34°C) blood cardioplegia. Patients at high risk for post cardiac surgery AKI identified using a modified risk score. [19] Exclusion criteria included: pre-existing inflammatory state (sepsis undergoing treatment, acute kidney injury within 5 days, chronic inflammatory disease or congestive heart failure), emergency or salvage procedure, ejection fraction <30%, critical preoperative state (Stage 3 AKI or requiring ionotropes, ventilation or intra-aortic balloon pump), or pregnancy.

### Blood sampling, storage and bias

Blood was collected pre-operatively, on return to the intensive care unit, and 6–12, 24, 48, 72 and 96 hours post-operatively. The total volume collected from each patient was 104.8 ml collected in clotting, EDTA, citrate and hirudin tubes (S-Monovette, Sarstedt, Nuembrecht, Germany) using venous lines, inserted as part of standard care during anaesthetic induction (perioperative samples) or by venipuncture. All anticoagulated samples were processed within 2 hours of collection and plasma (centrifuged twice at 1,500 × g) was frozen in single use aliquots at –80°C for subsequent analysis. Samples were identified by a trial acronym and the patient’s study ID. Routine measurements, including serum creatinine, were performed in NHS laboratories by personnel who were unaware that the participant was in a trial. For all other analyses, laboratory staff were blinded to the AKI status of patients.

### Leukocyte and platelets activation

were measured in citrated blood samples by flow cytometry (Cyan ADP, Beckman Coulter, Brea, CA). For platelets 5 μL of whole blood was incubated at RT for 25 min with 1:20 dilutions of either FITC-coupled PAC-1 (BD Biosciences), or CD62P. (Abcam, Cambridge, UK) together with PE-coupled CD41 (Affymetrix) antibodies in phosphate-buffered saline in a total volume of 100 µL. Red blood cells and leukocytes were gated out in forward and side scatter. For leukocyte analysis, red cells in 100 μL of whole blood were lysed with FACS Lysing Solution (BD Biosciences, Oxford, UK) for 10 min at RT. Leukocytes, gated in forward and side scatter, were labelled with FITC-coupled CD64, CD11b, CD16, CD14, and PE-coupled CD163 (Affymetrix, Santa Clara, CA) antibodies at 1:20 dilution for 25 min at room temperature (RT).

### Soluble biomarker analysis

Levels of soluble circulating biomarkers, CXCL1, IL-6, –8 and –10 and sICAM1 were measured by multiplex assays on the MAGPIX Analyser (BioTechne, USA).

### EV analysis

was performed in hirudinated plasma samples. Concentration and size distribution were estimated using NanoSight NS500, nanoparticle tracking device (Malvern Instruments, Malvern, United Kingdom) in citrated plasma samples. The flow cytometry analysis of EVs was performed with fluorescently labelled annexin V or antibodies against CD235a, CD41, CD144, CD142, CD14, CD16 CD284 and CD3. Fluorescently labelled isotype control IgGs were used as controls. All antibodies and isotype controls were from ThermoFisher. Plasma samples (20 μL) were incubated with with antibodies at 1:20 dilution in annexin V binding buffer (Thermo Fisher) for 25 minutes at room temperature (RT) in a total volume of 100 μL. The samples were analysed by flow cytometry (Cyan ADP; Beckman Coulter, Brea, CA). EV-specific gates (0.1 – 1µm) were determined using polystyrene particles (Spherotech, USA). Percentage of EVs positive for specific antibodies was adjusted for isotype controls and analysed as described in the statistical analysis section.

### MicroRNA analysis

RNA from EVs were isolated from citrated plasma using exoRNeasy kit (Qiagen) according to the manufacturer’s instructions. RNA (100 ng) was reverse transcribed using the Taq-Man Multiplex RT set for TaqMan Array Human MicroRNA Panel v2.0 (Applied Biosystems, Foster City, USA) and Veriti thermal cyclers. Pre-amplified complementary DNA from each pool was analysed using TaqMan Array Human MicroRNA Card A and B v2.0 (Applied Biosystems).

Quantitative PCR was carried out on an Applied BioSystems 7900HT thermocycler. All measurements were performed in triplicate. Any assays where the cycling threshold (Ct) value was lower than 20 were removed, and ΔCt (difference in cycling threshold) values were calculated (ΔCt = miRNA Ct– reference miRNA Ct) using reference control U6 snRNA-001973 present in duplicates on each miRNA array. Undetermined assays were assigned the maximum cycling value. Data were normalised across arrays using quantile normalisation. Differential expression analysis was performed with *limma* R package. [20]

Selected differentially expressed miRNAs were analysed using rotor-gene (Qiagen) quantitative real-time PCR instruments in samples from all patients using TaqMan Advanced miRNA assays for hsa-miR-17, hsa-miR-223, hsa-miR-486, hsa-miR-668, hsa-miRs-92a-1, hsa-miRs-920, hsa-miRs-518a-3p, hsa-miRs-133b and hsa-miRs-1262 (ThermoFisher) according to the manufacturer’s recommendations. The data was standardised against levels of hsa-miR-17 and hsa-miR-486, which showed constant average expression levels across all time-points and patient groups (**sFig. 1**).

**Figure 1.**
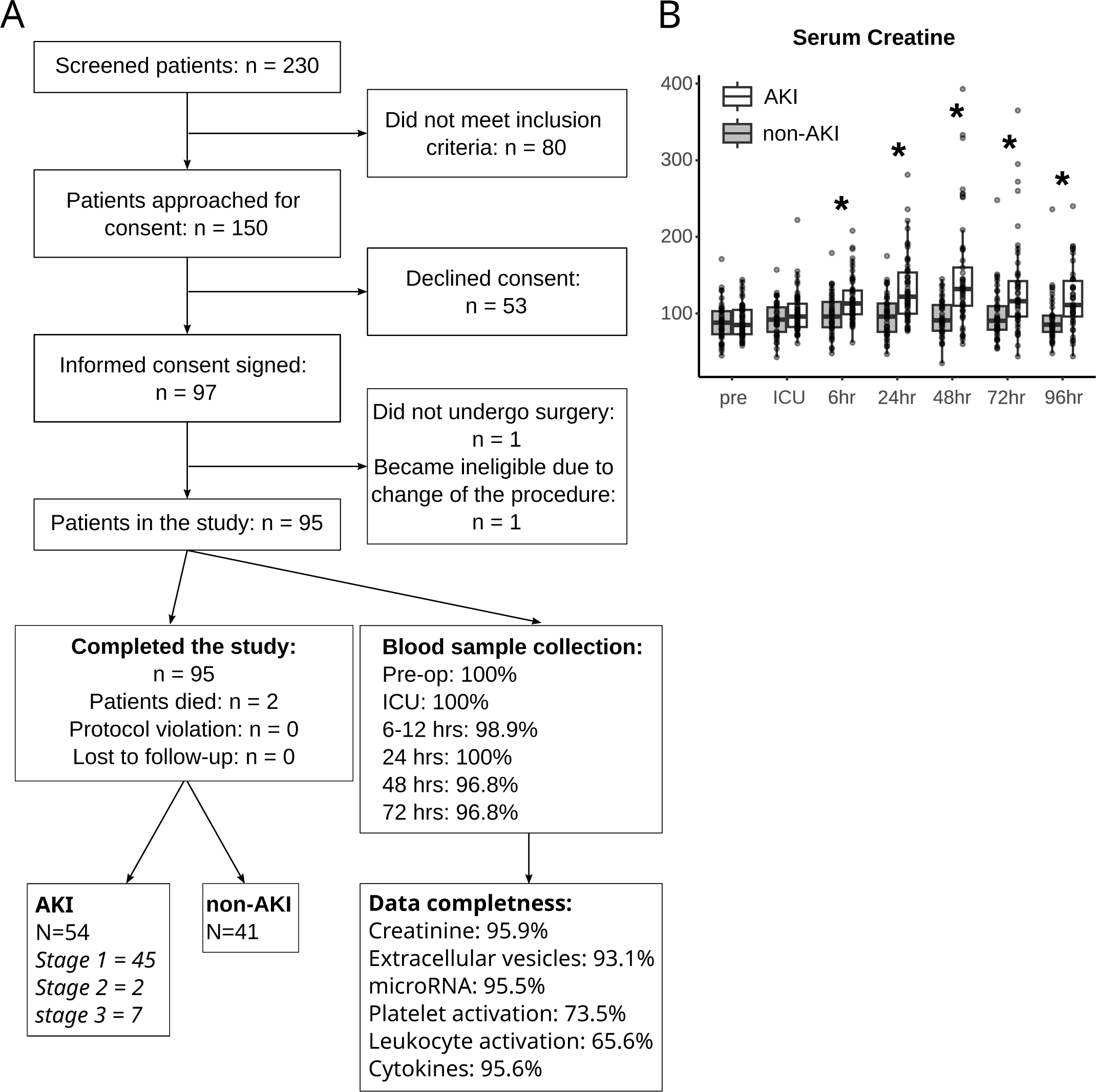
– (**A**) CONSORT diagram. (**B**) Levels of serum creatinine. Asterisks indicate a significant difference between no-AKI and AKI (*p*<0.05).

### Statistical analysis

was performed using R Studio Version 4.3.1 [21] and plots prepared with ggplot R package. [22] Data normality was assessed using the Shapiro-Wilk test, with normally distributed variables expressed as mean (± standard deviation [SD]) and non-normally distributed variables expressed as median (interquartile range [IQR]). Analyses were performed between patients with AKI and without AKI. Comparisons of miRNA analysed with PCR-arrays and EV size were adjusted for multiple comparisons using Benjamini-Hochberg method. Density plots were generated using *density* function, which calculates the distribution of a numeric variable using a kernel density estimates.

## Results

### Participants

Screening of 230 cardiac surgery patients identified 150 who were eligible for the study. Out of these, fifty-three declined consent and two became ineligible after giving consent. Ninety-five patients completed the study (**Fig. 1A**). Fifty-four of these patients (57%) developed post-cardiac surgery AKI, determined according to the Kidney Disease Improving Global Outcomes (KDIGO) definition, [23] with the majority (n=45) exhibiting stage 1, two patients developing stage 2, and 7 patients developing stage 3 AKI (**Fig. 1A**). Patient baseline demographics and intra-operative characteristics, shown in **Table 1**, were not different between the groups. The mean age was 72.8 (non-AKI, range 54-84) and 70.6 (AKI, range 50-84), with 85% in both groups being male. As expected, levels of serum creatinine were significantly higher in AKI patients 6 – 96 hours after surgery (**Fig. 1B**, **Table 1** and **eTable 1**).

**Table 1.** – Baseline and intraoperative characteristics. Tests between groups were conducted by exact test for categorical variables and t-test or non-parametric Kruskal–Wallis test for continuous variables. Data are presented as n (%) for categorical variables and mean (standard deviation, STD) or median (interquartile range) for continuous variables. Significant values are given in bold. * missing data: smoking staus (n=3), CCS (n=1), LVEF (n=5), left main steam diseased (n=4), Extent of coronary disease (n=4), Antiplatelets drugs (N=4)

|  | no AKI<br>(n = 41) | AKI<br>(n = 54) | p-value |
| --- | --- | --- | --- |
| Diabetes | 20 (48.8%) | 31 (57.4%) | 0.416 |
| CKD | 9 (22.0%) | 11 (20.4%) | 0.852 |
| Age (years) | 70.6 (6.9) | 72.8 (7.6) | 0.151 |
| Sex - Male | 35 (85.4%) | 46 (85.2%) | 0.980 |
| Caucasian | 41 (100.0%) | 52 (96.3%) | 0.504 |
| BMI | 31.7 (5.2) | 31.4 (6.1) | 0.745 |
| **Surgery type** |  |  |  |
| CABG | 19 (46.3%) | 19 (35.2%) | 0.324 |
| Valve | 15 (36.6%) | 19 (35.2%) |  |
| both/others | 7 (17.1%) | 16 (29.6%) |  |
| **Medical history** |  |  |  |
| Myocardial infarction | 7 (17.1%) | 10 (18.5%) | 0.856 |
| CVA/ TIA | 5 (12.2%) | 8 (14.8%) | 0.713 |
| Chronic pulmonary disease | 1 (2.4%) | 2 (3.7%) | 0.999 |
| Previous cardiac surgery | 2 (4.9%) | 4 (7.4%) | 0.696 |
| **Smoker** |  |  |  |
| Current | 12 (30.0%) | 22 (42.3%) | 0.478 |
| Ex-smoker | 18 (45.0%) | 19 (36.5%) |  |
| Never | 10 (25.0%) | 11 (21.2%) |  |
| **NYHA** |  |  |  |
| class I | 5 (12.2%) | 8 (14.8%) | 0.491 |
| class II | 29 (70.7%) | 32 (59.3%) |  |
| class III & IV | 7 (17.1%) | 14 (25.9%) |  |
| **CCS** |  |  |  |
| asymptomatic | 11 (27.5%) | 24 (44.4%) | 0.110 |
| class I | 10 (25.0%) | 11 (20.4%) |  |
| class II | 16 (40.0%) | 19 (35.2%) |  |
| class III | 3 (7.5%) | 0 (0.0%) |  |
| **Left ventricular ejection fraction (LVEF)** |  |  |  |
| Good(>49%) | 24 (63.2%) | 35 (67.3%) | 0.682 |
| Fair(30-49%) | 14 (36.8%) | 17 (32.7%) |  |
| **Left main stem diseased** | 4 (10.3%) | 9 (17.3%) | 0.342 |
| **Extent of coronary disease** |  |  |  |
| Normal | 6 (15.4%) | 11 (21.2%) | 0.287 |
| 1VD | 11 (28.2%) | 9 (17.3%) |  |
| 2VD | 8 (20.5%) | 6 (11.5%) |  |
| 3VD | 14 (35.9%) | 26 (50.0%) |  |
| **Anti-platelet drugs\*** |  |  |  |
| Aspirin (pre-operative) | 29 (74.4%) | 32 (61.5%) | 0.288 |
| P2Y12 inhibitors (stopped 5 days before surgery) | 5 (12.8) | 7 (13.5) | 1 |
| **Serum Troponin I** | 20 (15 -26) | 15 (15 -27) | 0.627 |
| **Serum creatinine (u mol/L)** | 88 (73 -103) | 85 (73 -105) | 0.848 |
| **Heart rate (bpm)** | 68.9 (12.4) | 66.4 (11.7) | 0.328 |
| **MAP (mmHg)** | 98.7 (11.9) | 97.5 (14.4) | 0.674 |
| **Hb (g/dL)** | 13.3 (1.7) | 12.9 (1.4) | 0.246 |
| **Hct (%)** | 39.1 (4.7) | 38.3 (4.0) | 0.418 |
| **WBC (x10<sup>9</sup>/L)** | 7.9 (1.8) | 8.1 (1.9) | 0.623 |
| **Platelets (x10<sup>9</sup>/L)** | 225.6 (76.8) | 214.9 (50.9) | 0.457 |
| **eGFR (ml/min/1.73m2)** | 82.4 (30.1) | 78.2 (22.4) | 0.453 |
| **Albumin (g/L)** | 40.5 (2.3) | 40.6 (3.6) | 0.825 |
| **Bilirubin (u mol/L)** | 13.1 (5.0) | 13.1 (6.5) | 0.992 |
| **AST (IU/L)** | 23.0 (4.8) | 25.1 (7.1) | 0.107 |
| **ALT (IU/L)** | 24.1 (11.0) | 24.8 (9.9) | 0.760 |
| **ALP (IU/L)** | 80.1 (26.5) | 83.1 (31.9) | 0.634 |
| **GGT (IU/L)** | 24 (18 -34) | 32 (19 -58) | 0.086 |
| **CBP time (min)** | 93 (82 -109) | 106 (82 -139) | 0.117 |
| **CrossClampTime (min)** | 55 (46 -77) | 65 (47 -97) | 0.259 |
| **Ventilation time (hr)** | 9 (7 -14) | 13 (10 -17) | **0.001** |
| **ICU stay (hr)** | 27 (25 -53) | 48 (25 -93) | 0.080 |
| **Hospital LOS (day)** | 8 (6 -9) | 10 (7 -13) | **0.017** |

The AKI patients had longer post-surgery ventilation times (AKI: 13, 10 –17 hours vs non-AKI 9, 7 – 14 hours, *p*=0.001) and longer hospital stays (AKI: 10, 7 –13 days vs 8, 6 –9 days, p=0.017).

### Platelet response

Aspirin (75mg per day) was given to 61 (64.2%) patients before surgery (32 [61.5%] AKI and 29 [74.4%] non-AKI). P2Y12 antagonists (Clopidogrel or Ticagrelor) were stopped five days before surgery. There was no significant difference in anti-platelet administration between AKI groups (**Table 1**).

The platelet response was analysed with a Multiplate aggregometer using ADP, ASPI and TRAP assays. Aspirin was the only drug that could affect platelet response during the study since patients received it daily. As expected, taking aspirin resulted in a lower AUC in the ASPI test, which was, and remained, below the cut-off value for aspirin responsiveness for the majority (78-88%) of aspirinated patients (**Fig. 2A**). There was no statistical difference in the ASPI in the numbers of aspirin high responders between AKI and non-AKI groups (**Fig. 2B**) or in responses to ASPI (**Fig. 2C**, **eTable 1**) or any time point.

**Figure 2.**
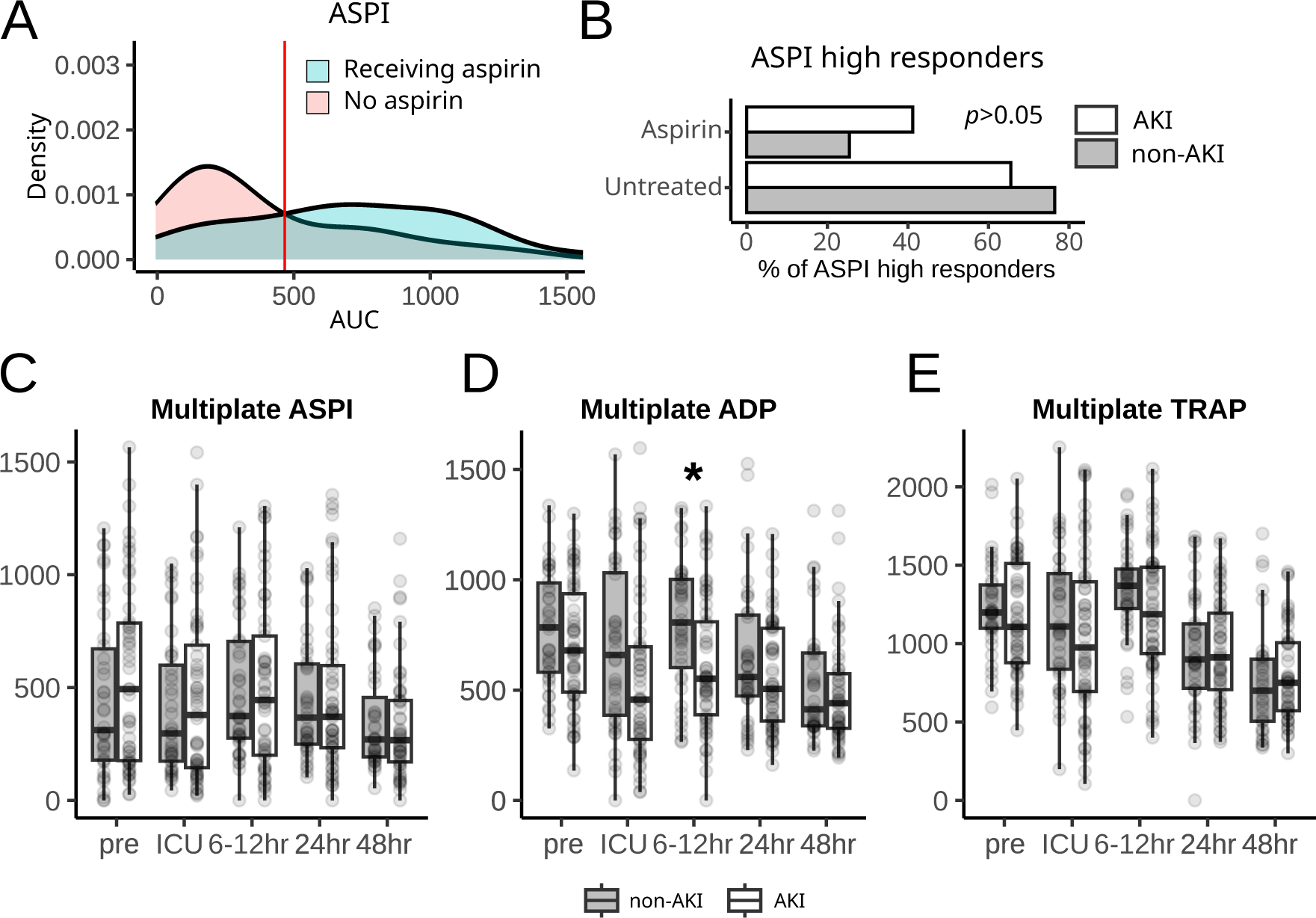
– (A) Density plot of the area under the curve (AUC) values acquired with Multiplate ASPI test. The vertical red line indicates the position of a cut-off value for ASPI high responders (AUC = 473). (B) Fraction of ASPI high responders in people with AKI or without. The analysis was performed for aspirin-treated and untreated patients. (C) Box plot of AUC data acquired using Multiplate ADP assay. Asterisks indicate a significant difference between AKI and non-AKI groups (p < 0.05). Open and grey boxes in B and D represent data from patients with and without AKI, respectively.

The range of responses to ADP was broad (**Fig. 2D**) and generally the ADP AUC was reduced following surgery. There was an overall trend for the AKI patients to have a lower ADP response at every time point, although the statistical difference between the groups was only observed at 6-12 hours after surgery (**Fig. 2D, eTable 1**). The TRAP test, which is insensitive to aspirin, indicated that the platelets showed the highest levels of aggregation 6-12 hours after surgery, which then fell over the subsequent 48 hours to levels that were lower than the responses before surgery. There was no difference in TRAP response between AKI groups (**Fig. 2E**, **eTable 1**).

### Thrombo-inflammatory cellular markers

Activation of blood cells implicated in the pathogenesis of AKI was measured using flow cytometry. These included markers of leukocyte activation (CD11b), monocyte activation (CD64 and CD163), and platelet activation (CD62P and activated GPIIb/IIIa), as well as circulating platelet-monocyte and platelet-granulocyte aggregates (CD14/CD41 and CD16/CD41, respectively).

Significantly higher levels of platelet activation (activated GPIIb/IIIa detected with PAC-1 antibody, **Fig. 3A** and **eTable 1**) were seen in the AKI patients 6-12 hours after surgery. P-selectin expression (CD41/CD62P), a marker of platelet degranulation, was no higher in the AKI patients, and did not change over time, but higher levels of granulocyte-platelet aggregates (a consequence of P-selectin expression on platelets) were seen in the AKI patients both before surgery and 6-12 and 24 hours afterwards (**Fig. 3B** and **eTable 1**).

**Figure 3.**
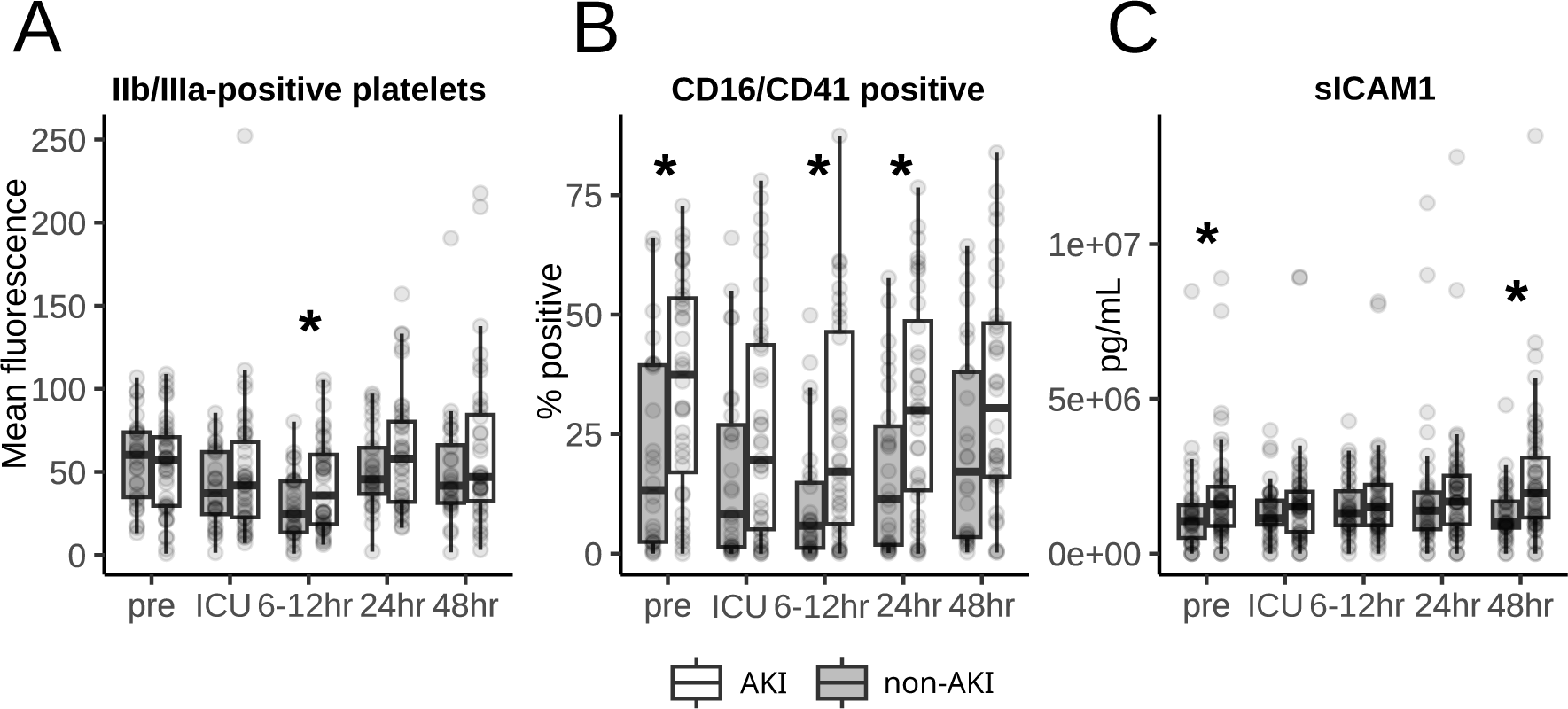
– (**A**) Box plot of mean expression values for IIb/IIIa detected with PAC-1 antibody in flow cytometry. (**B**) Box plot of a fraction of events positive for CD16 and CD41. (**C**) Box plot of sICAM1 concentrations in citrated plasma acquired with a MAGPIX device. Asterisks indicate a significant difference between AKI and non-AKI groups (p < 0.05). Open and grey boxes in A – C represent data from patients with and without AKI, respectively.

Analysis of soluble biomarkers of inflammation included CXCL1, soluble ICAM1 (sICAM1), IL-6, 8 and 10. Out of these, only sICAM1 was altered, and was significantly higher before surgery and 48 hours afterwards in the patients who developed AKI (**Fig. 3C** and **eTable 1**). Although there was no statistical difference in aspirin administration between the AKi groups before surgery, patients who received aspirin had significantly lower sICAM1 levels (1,27 µg/mL, IQR: 0.77-1.72 vs 1.62 µg/m, IQR: 0.96-2.71, *p*=0.046),

### Extracellular vesicles

EV concentrations ranged from 5.46 x 10^8^ – 14.56 x 10^8^ with no overall difference between AKI and non-AKI patients (**Fig. 4A, eTable 1**) or in particle sizes (**Fig. 4B**) at any time point. Derivation analysis identified EVs from monocytes (CD14), endothelial cells (CD144 and CD62E), granulocytes (CD16), red cells (CD235a), myeloid cells (CD284), T-cells (CD3) and platelets (CD41). We also assayed EVs positive for annexin V and tissue factor (CD142), which would indicate a pro-thrombotic phenotype. The most abundant EVs were of myeloid origin (CD284-positive; 8.77-15.88%) followed by annexin V-positive EVs (8.198-13.44%). The least abundant were EVs positive for CD41 (0.05-0.255%). However, only platelet-derived EVs (CD41-positive) were expressed at significantly higher levels in patients with AKI 24 hours after surgery (0.095%, 0.05-0.185 vs 0.05%, 0.02-0.18, *p*=0.028; **Fig. 4C** and **eTable 1**).

**Figure 4.**
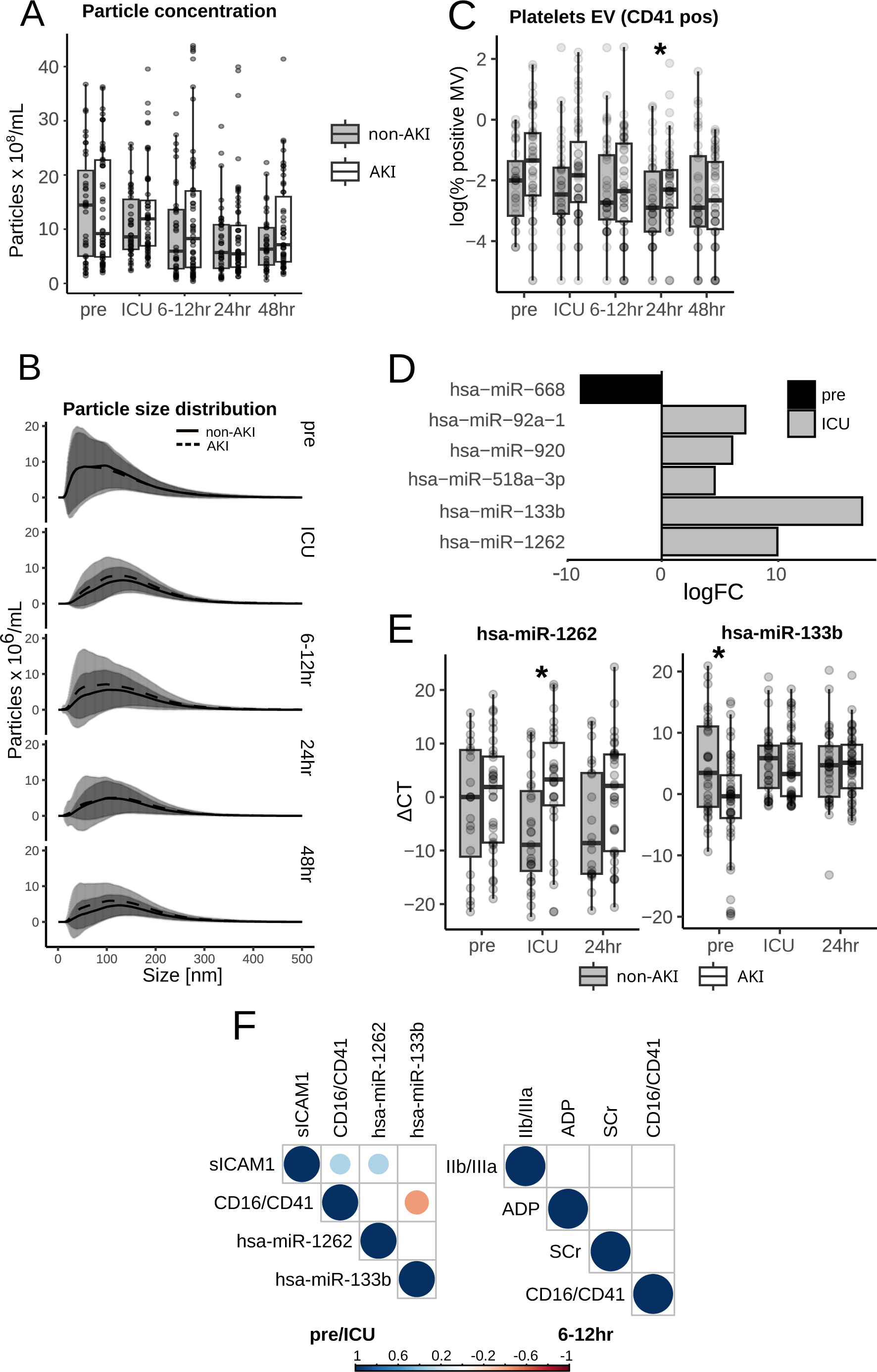
– (**A**) Total particle concentrations in citrated plasma. (**B**) Mean size distribution of particle concentrations (+/− SD). (**C**) Box plot of EVs positive for platelet marker CD41. (**D**) Bar plot showing the log fold difference in miRNA identified by qRT-PCR arrays between AKI and non-AKI. Only miRNA with adjusted *p*-value<0.05 are shown (see **eTable 2** for details). (**E**) Box plot of validation results for miR-133b and miR-1262. Asterisks indicate a significant difference between AKI and non-AKI groups (p < 0.05). Open and grey boxes in A, C and E represent data from patients with and without AKI, respectively. (**D**) Correlations (Spearman) between significant preoperative variables and ICU miR-1262, and between significant variables 6-12 hr after surgery. Blue circles indicate significant positive and red circles negative correlation.

### MicroRNAs

To identify AKI-related miRNA, 95 samples were pooled into 24 batches at each of three time points (pre, ICU and 24 hours after surgery) and analysed with qRT-PCR arrays for 754 human miRNA species. miR-668 was significantly lower in the AKI patients before surgery, while miRs-92a-1, 920, 518a-3p, 133b, and 1262 were higher in the samples from the AKI patients immediately after surgery (in ICU; **Fig, 4D** and **eTable 2**). Expression of these miRNAs was then verified by qRT-PCR in each of 95 samples at three time-points. Levels of miR-1262 were significantly higher immediately after surgery (ICU) in AKI patients, which was the result of lower expression levels in the non-AKI group. Levels of miR-133b were significantly lower in AKI patients before surgery (**Fig. 4E** and **eTable 1**)but not after surgery. Levels of miR-920 and mir-92a-1 were not different in AKI patients. Expression of miR-668 and miR-518a-3p were below detection level in most samples.

Out of all significant pre-operative variables, levels of platelet-granulocyte aggregates (CD16/CD41) correlated positively with levels of sICAM1 and negatively with levels of miR-133b. No significant correlation was observed between postoperative (6-12 hrs) variables (**Fig. 4F**).

### Sensitivity analysis in aspirin-treated patients

Given that most patients were receiving aspirin, we restricted the analysis to patients on aspirin only. The results of this analysis were very similar to those of all patients. Patients with AKI had higher levels of serum creatinine 6 – 96 hours after surgery, platelets-derived EV before surgery and at 24 hours post, levels miR-1262 at ICU, platelet-leukocyte aggregates before and 6-12 hours after surgery, and sICAM1 at 48 hours after surgery. Multiplate ADP AUC was lower at 6-12 hours after surgery in patients with AKI. However, we did not observe significant differences for activated GPIIb/IIIa and hsa-miR-133b, platelet-leukocyte aggregates 24 hours after surgery and sICAM1 before surgery (**eTable 3**).

## Discussion

Longitudinal analysis of cellular activation, inflammatory response, and levels of circulating EV and miRNA has shown a significant elevation of platelet activation, as demonstrated by platelets with activated GPIIb/IIIa and by platelet-granulocyte aggregates in cardiac surgery patients who develop AKI. Levels of platelet-granulocyte aggregates (CD16/CD41) were significantly higher pre-surgery and at 6-12 hours and 24 hours post-surgery in the AKI group. This group also had increased levels of platelets with activated GP IIb/IIIa (PAC-1 positive) and platelet-derived EVs at 6-12 and 24 hours post-surgery, respectively, alongside increased ADP insensitivity 6-12 hours after surgery.

Moreover, circulating sICAM1 levels were higher, and levels of plasma miR-133b were lower at baseline in the AKI group. Both of these correlated with levels of platelet-granulocyte aggregates at baseline. miR-1262 was most differentially expressed in the AKI group immediately after surgery and correlated with pre-operative sICAM1 levels.

The results demonstrate that platelet activation, resulting in platelet leukocyte aggregation are associated with AKI following cardiac surgery. Platelets have been shown to have an important role in experimental AKI, where platelet-granulocyte aggregates facilitate intraglomerular neutrophil recruitment and ROS production. [24] In rodents, platelets guide neutrophils to areas with reduced endothelial integrity like glomeruli, [3] whose glycocalyx is disrupted during AKI. [25] In the current study, sICAM1 and platelet leukocyte aggregates were positively correlated before surgery, suggesting that a pro-inflammatory state could prime the patients’ susceptibility to renal injury. miR-1262 has been associated with mesangial proliferative glomerulonephritis, [26] However, it remains to be tested whether its correlation with sICAM1 is functionally related. There is also little known about the role of miR-133b, which correlated negatively with platelet-granulocyte aggregates. It has been shown to regulate kidney fibrosis, by inhibiting connective tissue growth factor [27] and was previously associated with diabetic nephropathy, where its serum levels increase, [28] but no previous studies have associated miR-133b with either platelets or granulocytes. There are conflicting reports regarding miR-133 (unspecified strand or isoform), which was found enriched in human platelets by Diehl et al. [29] but not by Ambrose et al. [30]

Our data suggest that the observed desensitisation of platelets to ADP at 6-12 hours after surgery is not a consequence of the action of P2Y12 antagonists, which were not administered until 24 hours after surgery. It could be explained by the release of ADP in vivo from activated platelets, damaged red cells or other tissues. [31] We also show that aspirin did not affect the observed changes in platelet function in the AKI group. No trial to our knowledge has evaluated the renoprotective effect of anti-platelet agents in cardiac surgery. [32,33] Although, multiple RCTs have compared the effects of different anti-platelet agents on vein graft patency rates, arterial thromboses, and bleeding post-surgery, [34] none of these trials reported acute kidney injury event rates. Indirect evidence for a renoprotective effect of platelet inhibition comes from small trials of NO donors or prostacyclin [35–37] that have known anti-platelet effects and improve creatinine clearance after cardiac surgery. The increased levels of platelet-granulocyte aggregates in AKI patients before and after surgery brings into light the interaction between P-selectin and P-selectin glycoprotein ligand-1 (PSGL-1), which activated leucocytes facilitating leukocyte rolling and firm adhesion to the endothelium at sites of vascular injury or inflammation and promoting the release of pro-inflammatory cytokines and reactive oxygen species, thereby amplifying the inflammatory response. [38,39] Together, our results support the re-evaluation of existing trials or new trials of novel antiplatelet agents or blockers of P-selectin. These include drugs like crizanlizumab recently approved for sickle-cell disease [40] or Inclacumab, which is currently in the development phase.

As in any other study of AKI biomarkers, the major limitation is the use of serum creatinine to define renal injury. [41] More recent biomarkers, such as TIMP-2 and IGFBP7, have been shown to be useful diagnostic tools for AKI. However, they are not discriminatory in the cardiac surgery cohort. [11] Another limitation is the use of pooled samples for the initial identification of AKI-associated microRNAs. This was dictated by the budgetary limitations, and analysis of all patients, or a selected cohort of patients with well-defined clinical characteristics, could potentially result in the identification of more relevant microRNAs. Sample pooling can also be behind the discrepancies in miR-133b expression between discovery and validation. NanoSight, used to estimate particle concentration, does not discriminate between cell-derived, membrane-bound particles and other particulates in plasma such as lipoproteins. Therefore, we are not able to conclude whether the increases in specific particle sizes are associated with platelet-derived EVs. A final limitation is the small size of the study and high levels of missing data for flow cytometry analysis of platelet and leukocyte activation. We cannot be sure that a larger sample, or one with different eligibility criteria would not provide a different result.

## Authors’ contributions

Individual contributions to the study were as follows: GJM AHG and MJW designed the study, MJW and NB wrote the manuscript with critical input from GJM and AHG. LJ-D, TK and HA managed the conduct of the study. NS, BE-H, SS and KT collected samples and performed wet lab experiments. NB, NS, FYL and MJW performed statistical analyses

## Data Availability

All data produced in the present work are contained in the manuscript.

