## Supplementary tables and figures for "Acute kidney injury after cardiac surgery is associated with platelet activation"

### Figure S1

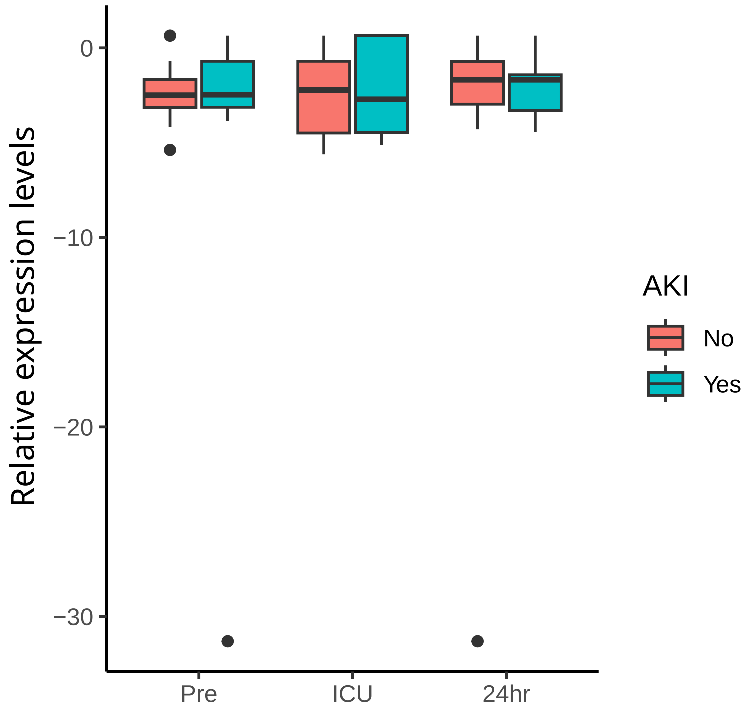


**Figure S1** - Boxplots of control miRNA used to normalise data across experiments. The values are processed sequencing reads, as described in the Methods section..

**eTable 1** – Summary of statistical analysis of all variables in the full dataset. Tests between groups were conducted by t-test or non-parametric Kruskal–Wallis test for continuous variables. Data are presented as mean (standard deviation, STD) or median (interquartile range) for continuous variables. Significant values are given in red.

| Type | Variable | Time | Normal distribution | AKI | non-AKI | p.value |
|---|---|---|---|---|---|---|
| AKI biomarkers | SCr | pre | No | 85 (73-104.75) | 88 (73-103) | 0.848 |
| AKI biomarkers | SCr | icu | No | 96 (82.25-112.75) | 92 (76-108) | 0.172 |
| AKI biomarkers | SCr | 6-12hr | No | 113 (99-130) | 96 (82-115) | 0.001 |
| AKI biomarkers | SCr | 24hr | No | 122 (99.75-153.5) | 96 (76-113) | <0.001 |
| AKI biomarkers | SCr | 48hr | No | 132 (110-160) | 91 (77.5-111) | <0.001 |
| AKI biomarkers | SCr | 72hr | No | 116 (96-142.25) | 90.5 (78.75-109.5) | <0.001 |
| AKI biomarkers | SCr | 96hr | No | 111 (96-142.5) | 85.5 (76-97.25) | <0.001 |
| EV analysis | Annexin V | pre | No | 10.348 (5.516-25.045) | 8.198 (4.853-14.263) | 0.263 |
| EV analysis | Annexin V | icu | No | 11.521 (7.352-17.58) | 11.508 (6.367-19.875) | 0.771 |
| EV analysis | Annexin V | 6-12hr | No | 10.765 (4.69-16.372) | 10.343 (5.889-18.661) | 0.504 |
| EV analysis | Annexin V | 24hr | No | 12.052 (8.602-22.61) | 13.144 (4.198-21.769) | 0.355 |
| EV analysis | Annexin V | 48hr | No | 10.809 (4.114-18.435) | 12.088 (6.875-18.915) | 0.489 |
| EV analysis | CD14 (monocytes) | pre | No | 2.375 (0.89-5.362) | 2.05 (0.785-5.94) | 0.769 |
| EV analysis | CD14 (monocytes) | icu | No | 1.805 (0.697-5.075) | 3.14 (1.24-5.03) | 0.242 |
| EV analysis | CD14 (monocytes) | 6-12hr | No | 1.66 (0.54-5.18) | 1.78 (0.968-4.718) | 0.297 |
| EV analysis | CD14 (monocytes) | 24hr | No | 2.11 (0.767-4.29) | 3.245 (0.965-5.707) | 0.242 |
| EV analysis | CD14 (monocytes) | 48hr | No | 2.21 (0.812-4.865) | 1.73 (0.815-4.11) | 0.698 |
| EV analysis | CD142 (tissue factor) | pre | No | 7.21 (2.458-15.31) | 5.77 (2.685-25.245) | 0.687 |
| EV analysis | CD142 (tissue factor) | icu | No | 5.905 (1.96-13.805) | 8.45 (5.21-14.97) | 0.127 |
| EV analysis | CD142 (tissue factor) | 6-12hr | No | 5.705 (1.97-9.82) | 7.875 (4.18-12.948) | 0.1 |
| EV analysis | CD142 (tissue factor) | 24hr | No | 4.72 (2.16-11.93) | 10.08 (3.643-19.898) | 0.194 |
| EV analysis | CD142 (tissue factor) | 48hr | No | 5 (2.035-15.682) | 6.32 (3.16-8.8) | 0.724 |
| EV analysis | CD144 (endothelial) | pre | No | 3.48 (1.785-8.56) | 2.865 (1.343-4.902) | 0.301 |
| EV analysis | CD144 (endothelial) | icu | No | 4.64 (2.085-12.79) | 4.86 (2.66-13.42) | 0.809 |
| EV analysis | CD144 (endothelial) | 6-12hr | No | 3.56 (2.04-7.95) | 6.285 (2.77-10.485) | 0.108 |
| EV analysis | CD144 (endothelial) | 24hr | No | 5.27 (2.09-9.37) | 5.71 (3.308-8.422) | 0.723 |
| EV analysis | CD144 (endothelial) | 48hr | No | 2.91 (1.21-6.293) | 3.65 (2.105-6.87) | 0.319 |
| EV analysis | CD16 (granulocytes) | pre | No | 3.64 (1.878-5.648) | 2.49 (1.4-4.217) | 0.186 |
| EV analysis | CD16 (granulocytes) | icu | No | 4.21 (2.57-7.232) | 3.76 (2.32-5.4) | 0.638 |
| EV analysis | CD16 (granulocytes) | 6-12hr | No | 4.77 (2.23-6.6) | 4.445 (2.475-7.033) | 0.702 |
| EV analysis | CD16 (granulocytes) | 24hr | No | 5.55 (2.62-7.54) | 4.285 (2.378-5.97) | 0.361 |
| EV analysis | CD16 (granulocytes) | 48hr | No | 6.245 (3.6-8.025) | 4.72 (3.26-6.35) | 0.132 |
| EV analysis | CD235a (red cells) | pre | No | 3.47 (1.208-10.887) | 2.645 (0.945-8.25) | 0.418 |
| EV analysis | CD235a (red cells) | icu | No | 2.35 (0.915-8.76) | 3.71 (1.92-6.1) | 0.554 |
| EV analysis | CD235a (red cells) | 6-12hr | No | 1.43 (0.61-4.61) | 3.115 (1.092-7.067) | 0.11 |
| EV analysis | CD235a (red cells) | 24hr | No | 1.94 (0.82-4.47) | 1.865 (0.627-5.352) | 0.819 |
| EV analysis | CD235a (red cells) | 48hr | No | 1.71 (0.597-5.092) | 2.05 (1.18-6.66) | 0.483 |
| EV analysis | CD284 (myeloid cells) | pre | No | 14.24 (8.02-24.81) | 13.525 (7.902-29.665) | 0.856 |
| EV analysis | CD284 (myeloid cells) | icu | No | 15.88 (4.47-25.787) | 14.51 (7.39-28.38) | 0.733 |
| EV analysis | CD284 (myeloid cells) | 6-12hr | No | 11.14 (7.47-17.92) | 15.87 (7.01-26.64) | 0.371 |
| EV analysis | CD284 (myeloid cells) | 24hr | No | 13.4 (5.1-31.51) | 12.865 (5.295-21.247) | 0.623 |
| EV analysis | CD284 (myeloid cells) | 48hr | No | 10.795 (4.192-22.282) | 8.77 (4.46-15.56) | 0.5 |
| EV analysis | CD3 (T-cells) | pre | No | 5.93 (3.143-13.482) | 7.135 (4.33-16.29) | 0.259 |
| EV analysis | CD3 (T-cells) | icu | No | 6.46 (3.267-12.688) | 7.88 (6.19-12.63) | 0.236 |
| EV analysis | CD3 (T-cells) | 6-12hr | No | 6.3 (3.74-10.19) | 7.8 (5.355-14.19) | 0.055 |
| EV analysis | CD3 (T-cells) | 24hr | No | 9.07 (5.32-14.613) | 8.48 (5.96-11.453) | 0.423 |
| EV analysis | CD3 (T-cells) | 48hr | No | 9.66 (4.093-22.778) | 6.88 (5.225-11.11) | 0.148 |
| EV analysis | CD41 (platelets) | pre | No | 0.255 (0.078-0.635) | 0.13 (0.038-0.252) | 0.05 |
| EV analysis | CD41 (platelets) | icu | No | 0.155 (0.062-0.483) | 0.08 (0.04-0.2) | 0.128 |
| EV analysis | CD41 (platelets) | 6-12hr | No | 0.09 (0.03-0.45) | 0.06 (0.032-0.305) | 0.769 |
| EV analysis | CD41 (platelets) | 24hr | No | 0.095 (0.05-0.185) | 0.05 (0.02-0.18) | 0.028 |
| EV analysis | CD41 (platelets) | 48hr | No | 0.065 (0.022-0.242) | 0.05 (0.025-0.295) | 0.986 |
| EV analysis | CD62E (endothelial) | pre | No | 2.46 (1.088-6.087) | 2.425 (1.55-4.64) | 0.41 |
| EV analysis | CD62E (endothelial) | icu | No | 2.635 (1.032-5.715) | 3.27 (1.77-5.12) | 0.617 |
| EV analysis | CD62E (endothelial) | 6-12hr | No | 2.7 (0.82-4.77) | 3.15 (1.885-6.738) | 0.176 |
| EV analysis | CD62E (endothelial) | 24hr | No | 4.085 (1.302-6.535) | 3.045 (1.24-4.452) | 0.182 |
| EV analysis | CD62E (endothelial) | 48hr | No | 3.51 (1.765-5.535) | 3.79 (1.56-4.665) | 0.453 |
| EV analysis | MV concentration | pre | No | 9.197 (4.887-22.753) | 14.455 (5.036-20.823) | 0.916 |
| EV analysis | MV concentration | icu | No | 11.915 (6.937-15.312) | 8.585 (6.281-15.496) | 0.225 |
| EV analysis | MV concentration | 6-12hr | No | 8.262 (2.955-17.048) | 5.952 (2.714-13.603) | 0.414 |
| EV analysis | MV concentration | 24hr | No | 5.458 (2.999-10.675) | 5.697 (2.784-10.822) | 0.806 |
| EV analysis | MV concentration | 48hr | No | 7.137 (3.987-15.998) | 6.323 (3.405-10.257) | 0.179 |
| miRNA | hsa-miR-1262 | pre | No | 8.281 (0-1986.495) | 1 (0-6714.537) | 0.566 |
| miRNA | hsa-miR-1262 | icu | No | 28.677 (0.241-26293.808) | 0 (0-4.311) | 0.005 |
| miRNA | hsa-miR-1262 | 24hr | No | 8.006 (0-2846.239) | 0 (0-88.788) | 0.106 |
| miRNA | hsa-miR-133b | pre | No | 0.694 (0.021-22.034) | 31.481 (0.126-61660.576) | 0.01 |
| miRNA | hsa-miR-133b | icu | No | 26.371 (0.72-3860.3) | 351.216 (2.716-2709.37) | 0.541 |
| miRNA | hsa-miR-133b | 24hr | No | 165.899 (2.676-3099.212) | 113.148 (0.846-2516.746) | 0.767 |
| miRNA | hsa-miR-920 | pre | No | 0 (0-0) | 0 (0-0.002) | 0.056 |
| miRNA | hsa-miR-920 | icu | No | 0 (0-0) | 0 (0-0) | 0.463 |
| miRNA | hsa-miR-920 | 24hr | No | 0 (0-0) | 0 (0-0) | 0.813 |
| miRNA | hsa-miR-92a-1 | pre | No | 0.009 (0-0.758) | 0.004 (0-1) | 0.733 |
| miRNA | hsa-miR-92a-1 | icu | No | 0.836 (0.008-9.821) | 0.007 (0-5.022) | 0.201 |
| miRNA | hsa-miR-92a-1 | 24hr | No | 0.785 (0-8.602) | 0 (0-1) | 0.064 |
| Multiplate analysis | ADP | pre | Yes | 703.615 (279.448) | 779.026 (262.799) | 0.194 |
| Multiplate analysis | ADP | icu | No | 457 (277-696) | 659.5 (385.75-1031.75) | 0.064 |
| Multiplate analysis | ADP | 6-12hr | Yes | 610.113 (301.135) | 797.000 (292.303) | 0.003 |
| Multiplate analysis | ADP | 24hr | No | 505.5 (359.75-780) | 559 (473-840) | 0.213 |
| Multiplate analysis | ADP | 48hr | No | 441 (327-574) | 413 (337.5-668) | 0.457 |
| Multiplate analysis | ASPI | pre | No | 493 (176-786) | 311.5 (178.25-671.25) | 0.312 |
| Multiplate analysis | ASPI | icu | No | 379 (145-688) | 296.5 (173.75-599.25) | 0.938 |
| Multiplate analysis | ASPI | 6-12hr | No | 445 (200-729) | 374 (275.5-704.25) | 0.822 |
| Multiplate analysis | ASPI | 24hr | No | 370.5 (233-597.5) | 367 (249-604.5) | 0.837 |
| Multiplate analysis | ASPI | 48hr | No | 267 (170-443) | 270 (192.5-456) | 0.663 |
| Multiplate analysis | TRAP | pre | Yes | 1168.962 (354.421) | 1227.921 (300.653) | 0.397 |
| Multiplate analysis | TRAP | icu | Yes | 1052.849 (526.757) | 1162.95 (433.772) | 0.272 |
| Multiplate analysis | TRAP | 6-12hr | Yes | 1211.396 (406.153) | 1342.4 (316.585) | 0.084 |

| | | | | | | |
|---|---|---|---|---|---|---|
| Multiplate analysis | TRAP | 24hr | Yes | 949.926 340.307732648069 | 947.816 (373.635) | 0.978 |
| Multiplate analysis | TRAP | 48hr | No | 750 (572-1005) | 700 (505-900.5) | 0.422 |
| Flow cytometry analysis | Activated GPIIb/IIIa (PAC-1) | pre | Yes | 53.766 (28.359) | 57.142 (25.778) | 0.605 |
| Flow cytometry analysis | Activated GPIIb/IIIa (PAC-1) | icu | No | 41.9 (22.72-68) | 37.195 (24.587-62.09) | 0.549 |
| Flow cytometry analysis | Activated GPIIb/IIIa (PAC-1) | 6-12hr | No | 35.885 (18.65-60.427) | 24.56 (13.65-44.435) | 0.043 |
| Flow cytometry analysis | Activated GPIIb/IIIa (PAC-1) | 24hr | No | 58.03 (32.03-80.318) | 45.695 (36.888-64.553) | 0.433 |
| Flow cytometry analysis | Activated GPIIb/IIIa (PAC-1) | 48hr | No | 46.93 (32.525-84.398) | 41.835 (31.363-66.2) | 0.398 |
| Flow cytometry analysis | CD11b | pre | No | 3.83 (0.76-12.015) | 2.28 (0.83-3.73) | 0.201 |
| Flow cytometry analysis | CD11b | icu | No | 8.28 (1.415-27.985) | 5.45 (0.472-21.317) | 0.372 |
| Flow cytometry analysis | CD11b | 6-12hr | No | 4.36 (0.82-18.69) | 3.645 (0.855-9.88) | 0.787 |
| Flow cytometry analysis | CD11b | 24hr | No | 6.31 (1.84-14.69) | 2.38 (0.89-10.367) | 0.251 |
| Flow cytometry analysis | CD11b | 48hr | No | 3.84 (1.19-21.173) | 1.42 (0.66-3.94) | 0.357 |
| Flow cytometry analysis | CD14/CD41 | pre | No | 7.245 (5.14-8.285) | 6.235 (4.784-7.755) | 0.294 |
| Flow cytometry analysis | CD14/CD41 | icu | No | 4.269 (2.783-5.969) | 3.33 (1.035-5.895) | 0.302 |
| Flow cytometry analysis | CD14/CD41 | 6-12hr | No | 7.875 (2.829-14.554) | 6.67 (1.376-11.7) | 0.301 |
| Flow cytometry analysis | CD14/CD41 | 24hr | No | 7.38 (3.74-13.081) | 4.7 (2.455-9.385) | 0.201 |
| Flow cytometry analysis | CD14/CD41 | 48hr | No | 7.52 (2.327-10.803) | 6.013 (2.596-11.314) | 0.971 |
| Flow cytometry analysis | CD16/CD41 | pre | No | 37.41 (16.993-53.438) | 13.304 (2.419-39.443) | 0.009 |
| Flow cytometry analysis | CD16/CD41 | icu | No | 19.682 (5.096-43.63) | 8.182 (1.396-26.928) | 0.133 |
| Flow cytometry analysis | CD16/CD41 | 6-12hr | No | 17.158 (6.192-46.396) | 5.823 (1.204-14.831) | 0.006 |
| Flow cytometry analysis | CD16/CD41 | 24hr | No | 29.968 (13.244-48.674) | 11.357 (1.816-26.631) | 0.019 |
| Flow cytometry analysis | CD16/CD41 | 48hr | No | 30.452 (16.14-48.169) | 17.124 (3.477-38.01) | 0.075 |
| Flow cytometry analysis | CD41/CD62P | pre | No | 11.14 (7.87-20.16) | 11.77 (9.06-18.94) | 0.583 |
| Flow cytometry analysis | CD41/CD62P | icu | No | 10.86 (8.14-18.13) | 10.12 (5.51-14.675) | 0.393 |
| Flow cytometry analysis | CD41/CD62P | 6-12hr | No | 10.07 (7.978-14.555) | 9.935 (6.522-13.608) | 0.572 |
| Flow cytometry analysis | CD41/CD62P | 24hr | No | 11.73 (7.387-15.41) | 9.82 (7.237-13.675) | 0.316 |
| Flow cytometry analysis | CD41/CD62P | 48hr | No | 12.34 (8.7-21.12) | 11.185 (5-16.415) | 0.259 |
| Flow cytometry analysis | CD64/CD163 | pre | No | 7.476 (5.805-10.05) | 6.79 (5.543-8.937) | 0.378 |
| Flow cytometry analysis | CD64/CD163 | icu | No | 8.75 (3.891-16.539) | 7.509 (1.975-12.405) | 0.308 |
| Flow cytometry analysis | CD64/CD163 | 6-12hr | No | 14.929 (6.672-28.555) | 11.061 (5.195-27.611) | 0.288 |
| Flow cytometry analysis | CD64/CD163 | 24hr | No | 16.21 (7.499-28.39) | 7.509 (5.081-16.515) | 0.05 |
| Flow cytometry analysis | CD64/CD163 | 48hr | No | 16.335 (7.939-30.59) | 8.913 (6.177-24.41) | 0.266 |
| Circulating biomarkers | CXCL1 | pre | No | 2.768 (2.318-10.4) | 3.199 (2.318-6.65) | 0.341 |
| Circulating biomarkers | CXCL1 | icu | No | 12.119 (2.462-30.367) | 6.679 (2.682-14.154) | 0.167 |
| Circulating biomarkers | CXCL1 | 6-12hr | No | 6.085 (2.318-16.255) | 3.989 (2.318-9.676) | 0.338 |
| Circulating biomarkers | CXCL1 | 24hr | No | 3.898 (2.318-13.169) | 2.461 (2.318-6.826) | 0.32 |
| Circulating biomarkers | CXCL1 | 48hr | No | 3.136 (2.318-14.852) | 2.61 (2.318-7.86) | 0.661 |
| Circulating biomarkers | ICAM1 | pre | No | 1599162.035 (888565.143-2159956.672) | 1050797.044 (503857.618-1563558.9) | 0.046 |
| Circulating biomarkers | ICAM1 | icu | No | 1513902.02 (696906.761-1998536.287) | 1141869.135 (934889.756-1717702.125) | 0.261 |
| Circulating biomarkers | ICAM1 | 6-12hr | No | 1491300.445 (911558.2-2225791.226) | 1309041.26 (911398.265-2018495.947) | 0.524 |
| Circulating biomarkers | ICAM1 | 24hr | No | 1679358.43 (939582.854-2518320.279) | 1385257.45 (787516.012-1977158.473) | 0.157 |
| Circulating biomarkers | ICAM1 | 48hr | No | 1948995.848 (1162919.005-3104333.25) | 1012597.923 (837179.132-1688613.032) | 0.002 |
| Circulating biomarkers | IL10 | pre | No | 0.355 (0.06-1.07) | 0.208 (0.06-0.937) | 0.561 |
| Circulating biomarkers | IL10 | icu | No | 23.377 (8.31-36.178) | 26.424 (10.154-59.832) | 0.222 |
| Circulating biomarkers | IL10 | 6-12hr | No | 8.146 (2.975-21.964) | 6.219 (2.32-16.935) | 0.528 |
| Circulating biomarkers | IL10 | 24hr | No | 3.085 (1.259-7.27) | 1.721 (0.3-6.29) | 0.172 |
| Circulating biomarkers | IL10 | 48hr | No | 1.311 (0.193-3.342) | 1.345 (0.06-3.7) | 0.745 |
| Circulating biomarkers | IL6 | pre | No | 0.52 (0.52-0.524) | 0.52 (0.52-0.52) | 0.581 |
| Circulating biomarkers | IL6 | icu | No | 53.786 (23.085-129.063) | 49.056 (25.734-79.706) | 0.535 |
| Circulating biomarkers | IL6 | 6-12hr | No | 45.864 (20.607-82.747) | 46.023 (23.639-60.262) | 0.742 |
| Circulating biomarkers | IL6 | 24hr | No | 34.397 (11.97-59.674) | 25.535 (13.926-45.318) | 0.542 |
| Circulating biomarkers | IL6 | 48hr | No | 28.154 (11.699-62.629) | 30.19 (14.471-74.294) | 0.939 |
| Circulating biomarkers | IL8 | pre | No | 0.003 (0.003-0.826) | 0.003 (0.003-0.957) | 0.744 |
| Circulating biomarkers | IL8 | icu | No | 3.138 (1.529-7.707) | 4.662 (1.67-6.969) | 0.692 |
| Circulating biomarkers | IL8 | 6-12hr | No | 3.492 (1.421-5.645) | 3.409 (1.794-5.447) | 0.939 |
| Circulating biomarkers | IL8 | 24hr | No | 2.082 (0.655-3.617) | 1.71 (0.425-3.757) | 0.709 |
| Circulating biomarkers | IL8 | 48hr | No | 1.848 (0.22-3.56) | 1.241 (0.238-2.709) | 0.438 |

**eTable 2 –** Significantly different miRs – logFC – log fold change; AveExpr – average expression; t – t-statistics; P.Value – unadjusted p-value; adj.P.Val – Benjamini-Hochberd-adjusted p-value, B – B-statistics

| Time | miRNA | logFC | AveExpr | t | P.Value | adj.P.Val | B |
|---|---|---|---|---|---|---|---|
| pre | has-miR-668 | -6.923 | -30.550 | -5.852 | <0.001 | <0.001 | 6.292 |
| ICU | hsa-miR-518a-3p | 4.541 | -31.097 | 210.544 | <0.001 | <0.001 | 114.872 |
| ICU | hsa-miR-133b | 17.189 | -26.748 | 5.342 | <0.001 | 0.001 | 4.461 |
| ICU | hsa-miR-92a | 7.172 | -30.102 | 4.933 | <0.001 | 0.002 | 3.005 |
| ICU | Hsa-miR-1262 | 9.919 | -8.115 | 4.487 | <0.001 | 0.007 | 1.583 |
| ICU | Hsa-miR-920 | 6.052 | -30.279 | 3.942 | <0.001 | 0.034 | -0.210 |

**eTable 3** – Summary of statistical analysis of all variables in patients receiving aspirin only. Tests between groups were conducted by t-test or non-parametric Kruskal–Wallis test for continuous variables. Data are presented as mean (standard deviation, STD) or median (interquartile range) for continuous variables. Significant values are given in red.

| Type | Variable | Time | Normal | AKI | non-AKI | p.value |
|---|---|---|---|---|---|---|
| AKI biomarkers | Serum creatinine | pre | Yes | 86.188 (19.42) | 86.793 (22.823) | 0.912 |
| AKI biomarkers | Serum creatinine | icu | No | 93 (80 - 103.75) | 88 (76 - 101) | 0.374 |
| AKI biomarkers | Serum creatinine | 6hr | No | 106 (96 - 123.5) | 94.5 (82 - 113.75) | 0.016 |
| AKI biomarkers | Serum creatinine | 24hr | No | 114 (97 - 141) | 100.5 (77.5 - 113) | 0.003 |
| AKI biomarkers | Serum creatinine | 48hr | No | 128.5 (106.25 - 142.5) | 91 (83 - 111) | 0.001 |
| AKI biomarkers | Serum creatinine | 72hr | No | 114 (91.5 - 133.5) | 91 (78 - 109) | 0.018 |
| AKI biomarkers | Serum creatinine | 96hr | No | 105.5 (89.5 - 131) | 85.5 (77.25 - 97.75) | 0.011 |
| EV analysis | Annexin V | pre | No | 11.828 (6.252 - 30.099) | 9.666 (5.183 - 14.799) | 0.201 |
| EV analysis | Annexin V | icu | No | 11.651 (8.997 - 19.112) | 13.585 (6.044 - 22.033) | 0.993 |
| EV analysis | Annexin V | 6hr | No | 11.135 (5.824 - 16.501) | 11.255 (6.344 - 19.241) | 0.636 |
| EV analysis | Annexin V | 24hr | No | 15.241 (9.366 - 24.579) | 11.471 (4.198 - 26.488) | 0.216 |
| EV analysis | Annexin V | 48hr | No | 9.477 (3.473 - 18.407) | 12.088 (9.572 - 18.727) | 0.279 |
| EV analysis | CD14 (monocytes) | pre | No | 3.98 (1.418 - 6.2) | 2.34 (0.785 - 7.98) | 0.946 |
| EV analysis | CD14 (monocytes) | icu | No | 1.92 (0.922 - 5.432) | 3.06 (1.255 - 6.365) | 0.506 |
| EV analysis | CD14 (monocytes) | 6hr | No | 1.76 (0.66 - 5.275) | 2.215 (1.102 - 5.228) | 0.329 |
| EV analysis | CD14 (monocytes) | 24hr | No | 2.335 (0.845 - 5.44) | 4.135 (1.465 - 5.905) | 0.325 |
| EV analysis | CD14 (monocytes) | 48hr | No | 2.65 (1.225 - 4.46) | 2.16 (1.29 - 5.79) | 0.941 |
| EV analysis | CD142 (tissue factor) | pre | No | 10.49 (3.896 - 19.472) | 12.27 (2.685 - 27.69) | 0.759 |
| EV analysis | CD142 (tissue factor) | icu | No | 8 (2.287 - 17.94) | 7.86 (4.828 - 14.15) | 0.755 |
| EV analysis | CD142 (tissue factor) | 6hr | No | 8.24 (2.185 - 10.78) | 7.875 (4.3 - 12.583) | 0.337 |
| EV analysis | CD142 (tissue factor) | 24hr | No | 6.28 (2.435 - 16.005) | 10.09 (4.715 - 20.123) | 0.429 |
| EV analysis | CD142 (tissue factor) | 48hr | No | 7.06 (2.93 - 19.935) | 6.58 (3.9 - 8.95) | 0.612 |
| EV analysis | CD144 (endothelial) | pre | No | 6.61 (2.645 - 11.08) | 2.975 (1.343 - 5.803) | 0.148 |
| EV analysis | CD144 (endothelial) | icu | No | 5.62 (2.783 - 13.555) | 4.245 (2.135 - 14.55) | 0.736 |
| EV analysis | CD144 (endothelial) | 6hr | No | 3.79 (2.325 - 9.755) | 6.94 (3.565 - 10.622) | 0.255 |
| EV analysis | CD144 (endothelial) | 24hr | No | 5.26 (2.208 - 10.375) | 5.98 (3.433 - 8.745) | 0.561 |
| EV analysis | CD144 (endothelial) | 48hr | No | 2.77 (1.585 - 6.18) | 3.7 (2.22 - 7.22) | 0.226 |
| EV analysis | CD16 (granulocytes) | pre | No | 3.64 (2.056 - 5.663) | 2.08 (1.298 - 3.848) | 0.108 |
| EV analysis | CD16 (granulocytes) | icu | No | 4.41 (2.587 - 6.207) | 3.005 (2.245 - 4.852) | 0.073 |
| EV analysis | CD16 (granulocytes) | 6hr | No | 4.92 (2.265 - 6.625) | 4.275 (2.725 - 6.365) | 0.894 |
| EV analysis | CD16 (granulocytes) | 24hr | No | 5.785 (3.788 - 7.883) | 4.375 (2.378 - 6.62) | 0.329 |
| EV analysis | CD16 (granulocytes) | 48hr | No | 6.75 (4.165 - 9.355) | 4.77 (2.45 - 6.64) | 0.06 |
| EV analysis | CD235a (red cells) | pre | No | 5.295 (1.388 - 15.085) | 2.8 (1.078 - 9.93) | 0.159 |
| EV analysis | CD235a (red cells) | icu | No | 3.99 (1.85 - 9.41) | 3.785 (1.622 - 5.897) | 0.742 |
| EV analysis | CD235a (red cells) | 6hr | No | 2.11 (0.74 - 5.355) | 2.79 (1.117 - 6.595) | 0.499 |
| EV analysis | CD235a (red cells) | 24hr | No | 2.345 (0.893 - 5.212) | 2.375 (0.977 - 5.457) | 0.994 |
| EV analysis | CD235a (red cells) | 48hr | No | 1.38 (0.355 - 4.065) | 2.68 (1.43 - 6.95) | 0.232 |
| EV analysis | CD284 (myeloid cells) | pre | No | 19.35 (10.46 - 25.325) | 15.05 (9.305 - 29.665) | 0.643 |
| EV analysis | CD284 (myeloid cells) | icu | No | 17.32 (8.015 - 33.469) | 15.37 (8.165 - 24.322) | 0.751 |
| EV analysis | CD284 (myeloid cells) | 6hr | No | 12.15 (5.035 - 19.92) | 16.135 (7.94 - 26.913) | 0.432 |
| EV analysis | CD284 (myeloid cells) | 24hr | No | 15.81 (4.845 - 35.127) | 11.005 (5.055 - 18.722) | 0.263 |
| EV analysis | CD284 (myeloid cells) | 48hr | No | 10.7 (1.81 - 19.53) | 8.77 (5.05 - 15.43) | 0.856 |
| EV analysis | CD3 (T-cells) | pre | No | 6.05 (3.235 - 13.698) | 9.505 (4.883 - 16.29) | 0.213 |
| EV analysis | CD3 (T-cells) | icu | No | 6.785 (3.845 - 11.373) | 7.86 (6.568 - 10.957) | 0.509 |
| EV analysis | CD3 (T-cells) | 6hr | No | 7.29 (4.065 - 10.495) | 8.745 (5.738 - 13.37) | 0.216 |
| EV analysis | CD3 (T-cells) | 24hr | No | 8.93 (5.92 - 14.33) | 8.57 (6.215 - 11.265) | 0.614 |
| EV analysis | CD3 (T-cells) | 48hr | No | 8.88 (4.9 - 15.445) | 7.59 (5.46 - 10.73) | 0.342 |
| EV analysis | CD41 (platelets) | pre | No | 0.355 (0.1 - 1.235) | 0.135 (0.045 - 0.37) | 0.023 |
| EV analysis | CD41 (platelets) | icu | No | 0.165 (0.07 - 0.723) | 0.06 (0.04 - 0.19) | 0.096 |
| EV analysis | CD41 (platelets) | 6hr | No | 0.11 (0.025 - 0.5) | 0.06 (0.04 - 0.328) | 0.643 |
| EV analysis | CD41 (platelets) | 24hr | No | 0.11 (0.062 - 0.238) | 0.05 (0.03 - 0.115) | 0.032 |
| EV analysis | CD41 (platelets) | 48hr | No | 0.06 (0.025 - 0.185) | 0.05 (0.03 - 0.28) | 0.791 |
| EV analysis | CD62E (endothelial) | pre | No | 2.22 (1.04 - 6.305) | 2.72 (1.617 - 5.315) | 0.278 |
| EV analysis | CD62E (endothelial) | icu | No | 3.13 (1.515 - 6.555) | 4.27 (2.077 - 5.3) | 0.813 |
| EV analysis | CD62E (endothelial) | 6hr | No | 3.27 (1.44 - 6.64) | 4.26 (2.277 - 7.778) | 0.402 |
| EV analysis | CD62E (endothelial) | 24hr | No | 4 (1.45 - 5.96) | 4.15 (2.13 - 4.985) | 0.743 |
| EV analysis | CD62E (endothelial) | 48hr | No | 3.65 (2.065 - 4.61) | 4.05 (2.52 - 4.84) | 0.76 |
| EV analysis | EV concentration | pre | No | 9.707 (4.696 - 21.838) | 15.02 (4.863 - 24.41) | 0.866 |
| EV analysis | EV concentration | icu | No | 10.533 (7.043 - 12.528) | 8.937 (6.355 - 15.465) | 0.719 |
| EV analysis | EV concentration | 6hr | No | 7.875 (2.694 - 16.305) | 6.1 (2.732 - 13.237) | 0.711 |
| EV analysis | EV concentration | 24hr | No | 5.677 (3.363 - 11.437) | 5.11 (3.367 - 10.08) | 0.486 |
| EV analysis | EV concentration | 48hr | No | 6.647 (3.498 - 15.153) | 6.17 (3.04 - 8.16) | 0.269 |
| miRNA | hsa-miR-1262 | pre | No | 27.328 (0-1700.292) | 0.509 (0-4883.161) | 0.677 |
| miRNA | hsa-miR-1262 | icu | No | 199.713 (14.878-167301.166) | 0.001 (0-4.311) | 0.016 |
| miRNA | hsa-miR-1262 | 24hr | No | 4.294 (0.001-3327.722) | 0 (0-299.063) | 0.159 |
| miRNA | hsa-miR-133b | pre | No | 0.694 (0.005-19.664) | 16.693 (0.063-25155.195) | 0.107 |
| miRNA | hsa-miR-133b | icu | No | 34.88 (1.629-26185.531) | 423.669 (9.207-3857.218) | 0.647 |
| miRNA | hsa-miR-133b | 24hr | No | 171.48 (6.089-2755.387) | 114.52 (2.97-1387.016) | 0.567 |
| miRNA | hsa-miR-920 | pre | No | 0 (0-0) | 0 (0-0.001) | 0.233 |
| miRNA | hsa-miR-920 | icu | No | 0 (0-0) | 0 (0-0) | 0.204 |
| miRNA | hsa-miR-920 | 24hr | No | 0 (0-0) | 0 (0-0) | 0.524 |
| miRNA | hsa-miR-92a-1 | pre | No | 0 (0-0.222) | 0.004 (0-0.052) | 0.596 |
| miRNA | hsa-miR-92a-1 | icu | No | 0.792 (0.024-22.092) | 0.01 (0-149.256) | 0.667 |
| miRNA | hsa-miR-92a-1 | 24hr | No | 1.136 (0-20.842) | 0 (0-11.398) | 0.173 |
| Multiplate analysis | ADP | pre | Yes | 665.25 (237.21) | 753.222 (283.415) | 0.207 |

| | | | | | | |
|---|---|---|---|---|---|---|
| Multiplate analysis | ADP | icu | No | 427 (223.5 - 658) | 548 (361 - 784) | 0.125 |
| Multiplate analysis | ADP | 6hr | No | 552 (475.5 - 867.5) | 817 (561 - 1002) | 0.025 |
| Multiplate analysis | ADP | 24hr | Yes | 549.829 (222.828) | 590.346 (265.239) | 0.531 |
| Multiplate analysis | ADP | 48hr | No | 362.5 (301.25 - 516) | 399 (335.25 - 602.5) | 0.336 |
| Multiplate analysis | ASPI | pre | No | 323 (165.5 - 772.5) | 286 (114.5 - 465.5) | 0.249 |
| Multiplate analysis | ASPI | icu | No | 251 (149 - 619.5) | 232 (137 - 493) | 0.599 |
| Multiplate analysis | ASPI | 6hr | No | 445 (198.5 - 623.5) | 365 (265 - 702) | 0.826 |
| Multiplate analysis | ASPI | 24hr | No | 310 (234 - 627) | 342 (265.25 - 455.75) | 0.983 |
| Multiplate analysis | ASPI | 48hr | No | 223.5 (146.75 - 348.5) | 225 (185.25 - 381.5) | 0.475 |
| Multiplate analysis | TRAP | pre | Yes | 1158.656 (350.609) | 1235.407 (324.391) | 0.387 |
| Multiplate analysis | TRAP | icu | Yes | 1060.548 (535.746) | 1115.552 (423.282) | 0.66 |
| Multiplate analysis | TRAP | 6hr | Yes | 1247.692 (392.467) | 1331.424 (302.441) | 0.311 |
| Multiplate analysis | TRAP | 24hr | Yes | 952.829 (273.392) | 842.2 (362.364) | 0.205 |
| Multiplate analysis | TRAP | 48hr | No | 673.5 (562.5 - 792.25) | 640 (504 - 846.25) | 0.628 |
| Flow cytometry analysis | Activated GPIIb/IIIa (PAC-1) | pre | Yes | 52.078 (29.695) | 56.848 (26.03) | 0.565 |
| Flow cytometry analysis | Activated GPIIb/IIIa (PAC-1) | icu | No | 45.84 (19.698 - 73.232) | 33.74 (16.72 - 51.78) | 0.293 |
| Flow cytometry analysis | Activated GPIIb/IIIa (PAC-1) | 6hr | No | 48.04 (18.43 - 60.562) | 26.78 (17.433 - 44.772) | 0.104 |
| Flow cytometry analysis | Activated GPIIb/IIIa (PAC-1) | 24hr | No | 50.28 (29.513 - 80.073) | 47.01 (34.18 - 56.56) | 0.71 |
| Flow cytometry analysis | Activated GPIIb/IIIa (PAC-1) | 48hr | No | 40.525 (25.315 - 85.828) | 43.965 (31.363 - 66.2) | 1 |
| Flow cytometry analysis | CD11b | pre | No | 3.715 (0.603 - 17.413) | 2.39 (0.6 - 4.685) | 0.582 |
| Flow cytometry analysis | CD11b | icu | No | 11.485 (0.34 - 29.6) | 9.46 (0.908 - 22.192) | 0.774 |
| Flow cytometry analysis | CD11b | 6hr | No | 1.76 (0.63 - 19.055) | 4.775 (0.727 - 13.085) | 0.864 |
| Flow cytometry analysis | CD11b | 24hr | No | 6.31 (1.31 - 9.735) | 2.51 (1.145 - 12.055) | 0.808 |
| Flow cytometry analysis | CD11b | 48hr | No | 3.8 (0.865 - 13.372) | 2.335 (0.672 - 3.99) | 0.79 |
| Flow cytometry analysis | CD14/CD41 | pre | No | 7.245 (6.036 - 9.147) | 6.35 (4.968 - 7.68) | 0.18 |
| Flow cytometry analysis | CD14/CD41 | icu | No | 4.269 (2.593 - 5.883) | 3.35 (1.15 - 5.853) | 0.434 |
| Flow cytometry analysis | CD14/CD41 | 6hr | No | 9.56 (2.703 - 15.674) | 5.5 (1.362 - 11.865) | 0.294 |
| Flow cytometry analysis | CD14/CD41 | 24hr | No | 7.764 (4.47 - 14.972) | 5.717 (2.191 - 12.03) | 0.32 |
| Flow cytometry analysis | CD14/CD41 | 48hr | No | 7.852 (2.27 - 9.811) | 6.265 (2.995 - 15.774) | 0.727 |
| Flow cytometry analysis | CD16/CD41 | pre | No | 42.801 (20.605 - 52.947) | 11.936 (2.44 - 39.315) | 0.003 |
| Flow cytometry analysis | CD16/CD41 | icu | No | 21.366 (4.58 - 39.51) | 7.872 (1.689 - 26.794) | 0.227 |
| Flow cytometry analysis | CD16/CD41 | 6hr | No | 19.569 (8.271 - 48.52) | 4.781 (1.158 - 13.887) | 0.014 |
| Flow cytometry analysis | CD16/CD41 | 24hr | No | 27.988 (10.389 - 38.298) | 12.24 (2.051 - 30.151) | 0.155 |
| Flow cytometry analysis | CD16/CD41 | 48hr | No | 30.452 (16.12 - 47.194) | 23.408 (3.642 - 46.02) | 0.357 |
| Flow cytometry analysis | CD41/CD62P | pre | No | 11.77 (8.123 - 24.302) | 13.365 (9.012 - 18.95) | 0.801 |
| Flow cytometry analysis | CD41/CD62P | icu | No | 11.46 (8.515 - 18.23) | 10.02 (4.615 - 13.89) | 0.17 |
| Flow cytometry analysis | CD41/CD62P | 6hr | No | 10.925 (8.512 - 16.135) | 10.62 (7.01 - 13.355) | 0.504 |
| Flow cytometry analysis | CD41/CD62P | 24hr | No | 12.34 (8.54 - 14.73) | 10.11 (7.38 - 14.27) | 0.741 |
| Flow cytometry analysis | CD41/CD62P | 48hr | No | 12.34 (8.765 - 18.7) | 10.23 (4.69 - 16.22) | 0.316 |
| Flow cytometry analysis | CD64/CD163 | pre | No | 7.238 (5.795 - 10.135) | 6.77 (5.684 - 8.748) | 0.493 |
| Flow cytometry analysis | CD64/CD163 | icu | No | 5.359 (3.463 - 15.035) | 7.614 (1.892 - 12.404) | 0.745 |
| Flow cytometry analysis | CD64/CD163 | 6hr | No | 14.89 (7.224 - 34.879) | 10.085 (4.702 - 20.658) | 0.218 |
| Flow cytometry analysis | CD64/CD163 | 24hr | No | 15.659 (6.395 - 24.184) | 8.527 (4.57 - 16.269) | 0.238 |
| Flow cytometry analysis | CD64/CD163 | 48hr | No | 17.861 (7.676 - 29.341) | 9.416 (6.618 - 21.383) | 0.53 |
| Circulating biomarkers | CXCL1 | pre | No | 3.744 (2.318 - 10.585) | 3.199 (2.318 - 6.647) | 0.269 |
| Circulating biomarkers | CXCL1 | icu | No | 11.112 (2.615 - 34.176) | 6.712 (3.332 - 13.968) | 0.244 |
| Circulating biomarkers | CXCL1 | 6hr | No | 6.567 (2.318 - 15.784) | 4.911 (2.318 - 9.637) | 0.313 |
| Circulating biomarkers | CXCL1 | 24hr | No | 5.727 (2.318 - 15.051) | 2.318 (2.318 - 6.65) | 0.266 |
| Circulating biomarkers | CXCL1 | 48hr | No | 2.768 (2.318 - 17.557) | 2.349 (2.318 - 6.65) | 0.475 |
| Circulating biomarkers | ICAM1 | pre | No | 1308557.275 (839981.001 - 1732249.146 | 1197292.905 (543823.482 - 1532129.496 | 0.269 |
| Circulating biomarkers | ICAM1 | icu | No | 1497879.046 (647305.841 - 2084625.561 | 1095508.272 (806047.016 - 1643080.197 | 0.425 |
| Circulating biomarkers | ICAM1 | 6hr | No | 1298714.983 (829995.511 - 2210903.846 | 1304970.189 (813370.504 - 2149794.037 | 0.921 |
| Circulating biomarkers | ICAM1 | 24hr | No | 1654346.776 (779856.163 - 2549254.186 | 1465262.722 (852929.49 - 2208904.562) | 0.845 |
| Circulating biomarkers | ICAM1 | 48hr | No | 1750900.171 (1052195.998 - 3057463.00 | 1115578.167 (784847.679 - 1668053.479 | 0.035 |
| Circulating biomarkers | IL10 | pre | No | 0.06 (0.06 - 0.922) | 0.481 (0.06 - 1.074) | 0.553 |
| Circulating biomarkers | IL10 | icu | No | 23.207 (9.614 - 35.575) | 36.22 (10.347 - 58.5) | 0.157 |
| Circulating biomarkers | IL10 | 6hr | No | 8.049 (3.017 - 21.394) | 6.421 (2.395 - 17.51) | 0.803 |
| Circulating biomarkers | IL10 | 24hr | No | 5.028 (1.53 - 7.69) | 2.365 (0.472 - 6.31) | 0.197 |
| Circulating biomarkers | IL10 | 48hr | No | 1.548 (0.528 - 3.195) | 1.026 (0.06 - 2.627) | 0.319 |
| Circulating biomarkers | IL6 | pre | No | 0.52 (0.52 - 0.521) | 0.52 (0.52 - 0.52) | 0.712 |
| Circulating biomarkers | IL6 | icu | No | 52.094 (21.971 - 129.317) | 43.429 (26.151 - 85.071) | 0.681 |
| Circulating biomarkers | IL6 | 6hr | No | 46.908 (19.698 - 85.864) | 52.963 (21.065 - 60.958) | 0.825 |
| Circulating biomarkers | IL6 | 24hr | No | 46.319 (12.592 - 74.567) | 27.611 (18.354 - 45.902) | 0.38 |
| Circulating biomarkers | IL6 | 48hr | No | 29.903 (16.168 - 64.577) | 30.337 (14.471 - 74.294) | 0.779 |
| Circulating biomarkers | IL8 | pre | No | 0.003 (0.003 - 0.386) | 0.003 (0.003 - 0.807) | 0.662 |
| Circulating biomarkers | IL8 | icu | No | 2.839 (1.252 - 8.194) | 5.291 (1.19 - 7.24) | 0.656 |
| Circulating biomarkers | IL8 | 6hr | No | 3.552 (1.397 - 5.67) | 3.79 (2.095 - 6.285) | 0.693 |
| Circulating biomarkers | IL8 | 24hr | No | 2.003 (0.442 - 3.646) | 1.83 (0.416 - 3.449) | 0.797 |
| Circulating biomarkers | IL8 | 48hr | No | 1.326 (0.112 - 3.035) | 1.326 (0.386 - 2.265) | 0.628 |
